# COVID-19 propagation by diffusion - a two-dimensional approach for Germany

**DOI:** 10.1101/2021.01.15.21249893

**Authors:** Günter Bärwolff

## Abstract

Diffusion comes anytime and everywhere. If there is a gradient or a potential difference of a quantity a diffusion process happens and this ends if an equilibrium is reached only. The concentration of a species maybe such quantity, or the voltage. An electric currant will be driven by a voltage difference for example.

In this COVID-19 pandemic one observes both regions with low incidence and other ones with high incidence. The local different people density could be a reason for that. In populous areas like big cities or congested urban areas higher COVID-19 incidences could be observed than in rural regions.

The aim of this paper consists in the application of a diffusion concept to describe one possible issue of the the COVID-19 propagation.

This will be discussed for the German situation based on the quite different incidence data for the different federal states of Germany.

With this ansatz some phenomenoms of the actual development of the pandemic could be confirmed. The model gives a possibility to investigate certain scenarios like border-crossings or local spreading events and their influence on the COVID-19 propagation as well.

## 1 Introduction and the mathematical model

The mathematical modeling of COVID-19 with SIR-type models ([5], [6]) leads to averaged results and does not take unequal peopling or populousness into account. But it is well known, that these issues play an important role in the local pandemic evolution. With the consideration of local-dependent density of people and a diffusion model we try to resolve the COVID-19 propagation in a finer manner.

What is a good choice of a quantity to describe the COVID-19 spread? The WHO and national health institutions measure the COVID-19 spread with the seven-days-incidence (sometimes also the fourteen-days incidence) of infected people per 100000 inhabitants. In Germany it is possible to control or trace the history of infected people by local health institutions if the seven-days incidence has a value less than 50. But at the end of December 2020 and the begin of January 2021 the averaged incidence is about 140, and in some hotspot federal states like Saxony greater than 300. If the social and economical life should be sustained there are several possibilities to transmit the COVID-19 virus anyhow. The following ones should be mentioned:

- Commuters and employers on the way to there office or to there position of employment especially including medical and nursing staff.
- Pupils and teachers in schools and on the way to school and in the school.
- People buying every day necessities using shopping centers.
- Postmen, suppliers and deliverers.

All these activities take place during the so called lockdown in Germany with the result of ongoing propagation of the pandemic. Also the unavailable center of power in the decentralized federal state Germany. This leads often to solo efforts of some federal states.

From authoritarian countries like China or Singapore with a quite different civilization and other cultural traditions than the German ones for example it is known, that the virus propagation could be stopped with very rigorous measures like the strict prohibition of the social and economic life (see 2^1^), i.e. activities which mentioned above are absolutely forbidden.

This is inconceivable in countries like Germany, Austria, the Netherlands or other so called democratic states with a western understanding of freedom and self-determination (see 1). But in the consequences of such a western life style they have to live with a more or less consecutive activity of the COVID-19 pandemic. And that is the reason for the following trial to describe one aspect of the pandemic by a diffusion model. In another context a similar model was discussed in [4].

In the following diffusion concept the seven-days incidence should be denoted by *s* and *s* should serve as the quantity which will be influenced by it’s gradients between different levels of incidence in the federal states of Germany. At the the mathematical model of diffusion leads to a partial differential equation for the considered quantity (here *s*)

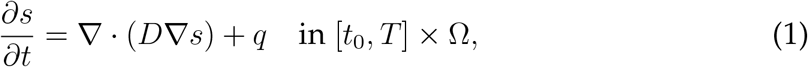

where Ω ⊂ ℝ^2^ is the region which will be investigated, for example the national territory of Germany, *D* is a diffusion coefficient, depending on the locality *x* ∈ Ω, [*t*_0_, *T*] is the time interval of interest, and *q* is a term which describes sources or sinks of possible infections.

Beside the equation (1) one needs initial conditions for *s*, for example

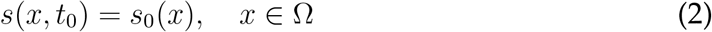

and boundary conditions

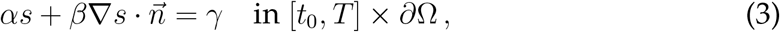

where *α, β* and *γ* are real coefficients and by *∂*Ω =: Γ the boundary of the region Ω will be denoted. 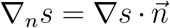 is the directional derivative of *s* in the direction of the outer normal vector 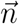 on Γ. The choice of *α* = 0, *β* = 1 and *γ* = 0 for example leads to the homogeneous Neumann boundary condition

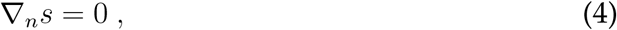

which means no import of *s* at the boundary Γ. In other words, (4) describes closed borders to surrounding countries outside Ω.

The diffusion coefficient function *D* : Ω → ℝ is responsible for the intensity or velocity of the diffusion process. From fluid or gas dynamics one knows from [1] the formula

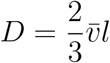

with the averaged particle velocity 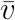 and the mean free path *l*. The application of this ansatz to the movement of people in certain areas requires assumptions for 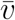 and *l*. If we consider a circular or quadratic region with the area *A* and a number of inhabitants *N* who are distributed equally *l* could be approached by

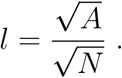

For the velocity 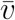 we assume 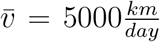 ([3]). Because of the different areas and numbers of inhabitants of the federal states of Germany *D* will be a local depending non-constant function.

If there are no sources or sinks for *s*, i.e. *q* = 0, and the borders are closed which means the boundary condition (4), the initial boundary value problem (1), (2), (4) has the steady state solution

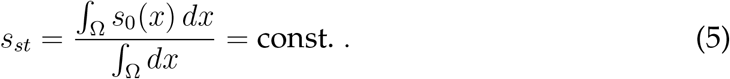

This is easy to verify and this property is characteristic for diffusion processes which tend to an equilibrium. It is quite complicated to model the source-sink function *q* in an appropriate kind. *q* depends of the behavior of the population and the health policy of the different federal states. It’s only possible to work with very coarse guesses. It is known that the people in Schleswig-Holstein is exemplary with respect to the recommendations to avoid infection with the COVID-19 virus and this means *q <* 0. On the other hand it is known from Saxony the many people belief there is not a jot of truth in the pandemic, which means *q >* 0 for a long time (now the government of Saxony changed the policy which leads to *q <* 0).

But regardless of these uncertainties one can get information about the pandemic propagation for example the influence of hotspots of high incidences (Saxony) to regions with low incidences (South of Brandenburg) for example.

## 2 Data of the different federal states of Germany

At the beginning of the year 2021 (14th of January) the Robert-Koch-Institut which is responsible for the daily COVID-19 data collection published the seven-days incidence data (of January the 14th, 2021, see [2]) summarized in table 1.

**Table 1:** 7-days incidence, people density [*/km*^2^], inhibitants [*/*100000], area of the federal states of Germany [*km*^2^]

| states | 7-days incidence | density | inhabitants | area |
| --- | --- | --- | --- | --- |
| Schleswig-Holstein | 92 | 183 | 2904 | 15804 |
| Hamburg | 115 | 2438 | 1847 | 755 |
| Mecklenburg-West Pomerania | 117 | 69 | 1608 | 23295 |
| Lower Saxony | 100 | 167 | 7994 | 47710 |
| Brandenburg | 212 | 85 | 2522 | 29654 |
| Berlin | 180 | 4090 | 3669 | 891 |
| Bremen | 84 | 1629 | 681 | 419 |
| Saxony-Anhalt | 241 | 109 | 2195 | 20454 |
| Thuringia | 310 | 132 | 2133 | 16202 |
| Saxony | 292 | 221 | 4072 | 18450 |
| Bavaria | 160 | 185 | 13125 | 70542 |
| Baden-Wuerttemberg | 133 | 310 | 11100 | 35784 |
| North Rhine-Westphalia | 131 | 526 | 17947 | 34112 |
| Hesse | 141 | 297 | 6288 | 21116 |
| Saarland | 160 | 385 | 987 | 2571 |
| Rhineland-Palatinate | 122 | 206 | 4094 | 19858 |
| Munic | 156 | 4700 | 1540 | 310 |

The values of table 1 are used as initial data for the function *s*_0_ of (2).

As a base for the determination of the diffusion coefficient function we use the data of table 1.

The unit of the diffusion function *D* will be [*km*^2^*/day*]. Eventual sources or sinks will be gauged in [*/day*]. The incidence *s* is dimensionless.

## 3 The numerical solution of the initial boundary value problem (1),(2),(4)

Based on the subdivision of Ω (area of Germany) into finite rectangular cells *ω*_*j*_, *j* ∈ *I*_Ω_ and 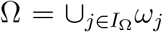 the equation (1) will be spatial discretized with a finite volume method (*I*_Ω_ is the index set of the finite volume cells). Together with the discrete boundary condition (4) we get a semi-discrete system continuous in time

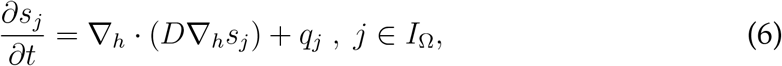

where the index *h* means the discrete versions of the ∇-operator.

The time discretization is done with an implicit Euler scheme. This allows us to work without strict restrictions for the choice of the discrete time-step Δ_*t*_. At every time-level it is to solve the linear equation system

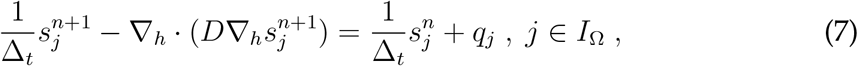

for 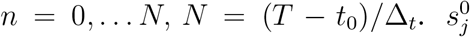 was set to the incidence *s*_0_(*x*) for *x* ∈ *ω*_*j*_, *j* = 1, …, *I*_Ω_. The solution of equation system (7) for a certain time-level *n* is done with an iterative method.

## 4 Numerical simulation results

In fig. 4 the region Ω is adumbrated. The size of finite volume cells is Δ_*x*_ × Δ_*y*_ = (8*km* × 8*km*). First we start with the case *q* = 0. Δ_*t*_ was set to one day. In the following figs. 5-10 the initial state and the results of the diffusion process for the development of the seven-days incidence after 50 and 120 time-steps (days) are shown. The left figs. show a view from west to east, and the right figs. show the view from north to south. The initial state is a piece-wise constant function with values of the seven-days incidence of the 16 federal states where we consider munic as a town with over a million inhabitants separately (it was excluded from Bavaria). Especially in the border regions (Saxony - Brandenburg, Saxony - Bavaria, Saxony - Thuringia) one can observe a transfer of incidence from the high level incidence of Saxony to the neighbored federal states. Also the high incidence level of Berlin was transferred to the surrounding area of Brandenburg. The north states with a low incidence level were only influenced by the other states weakly.

**Figure 1:**
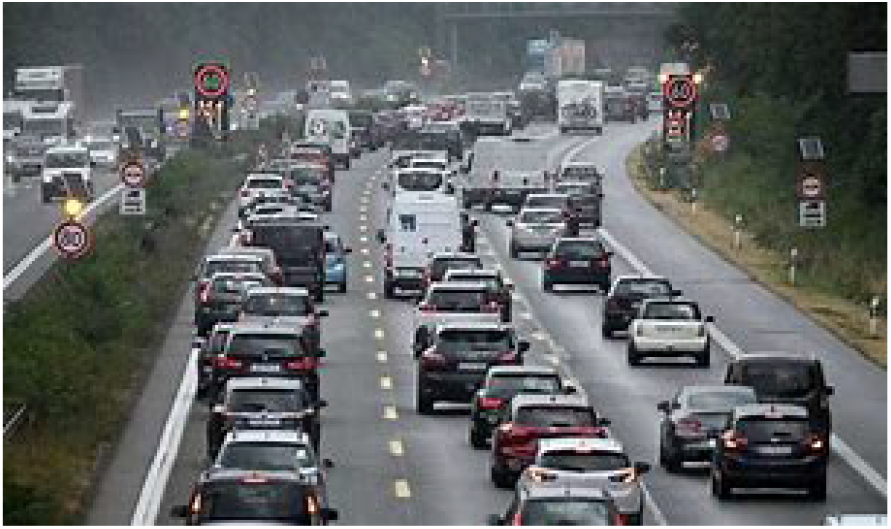
German lockdown

**Figure 2:**
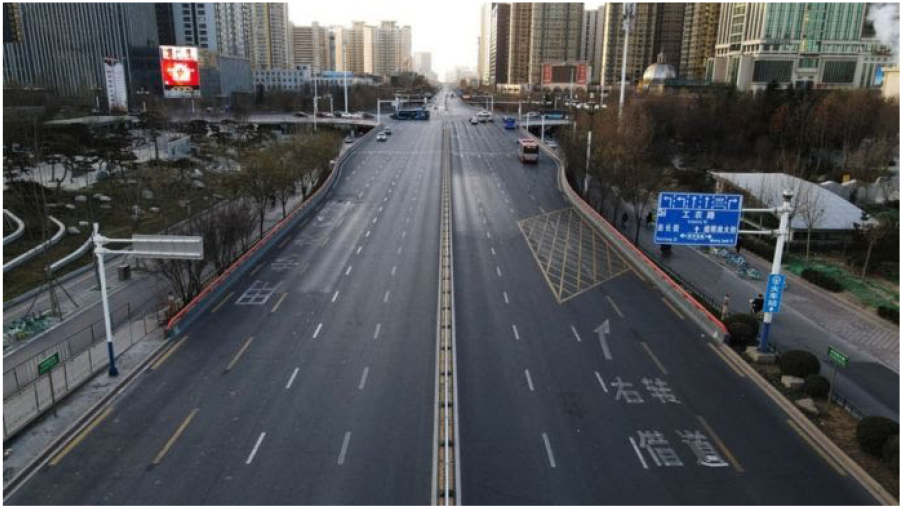
Chinese lockdown

**Figure 3:**
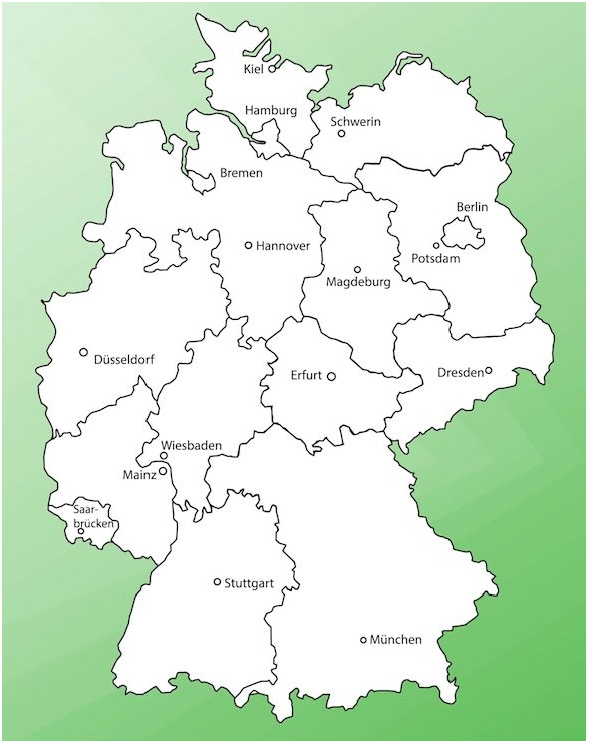
Germany map

**Figure 4:**
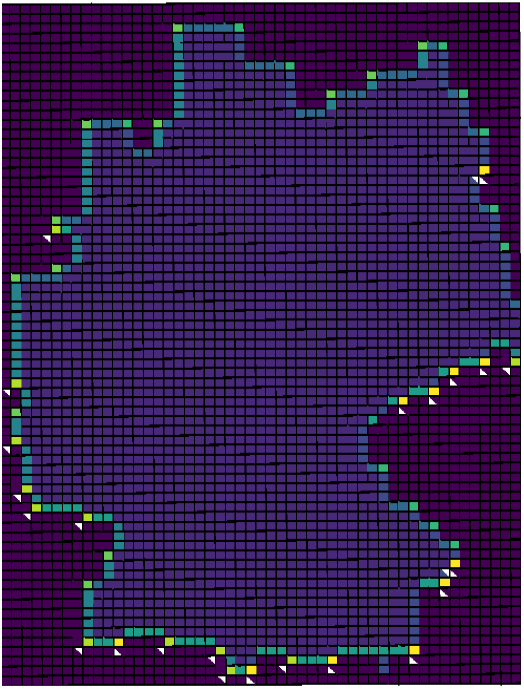
Coarse contour of Ω and it’s discretization

**Figure 5:**
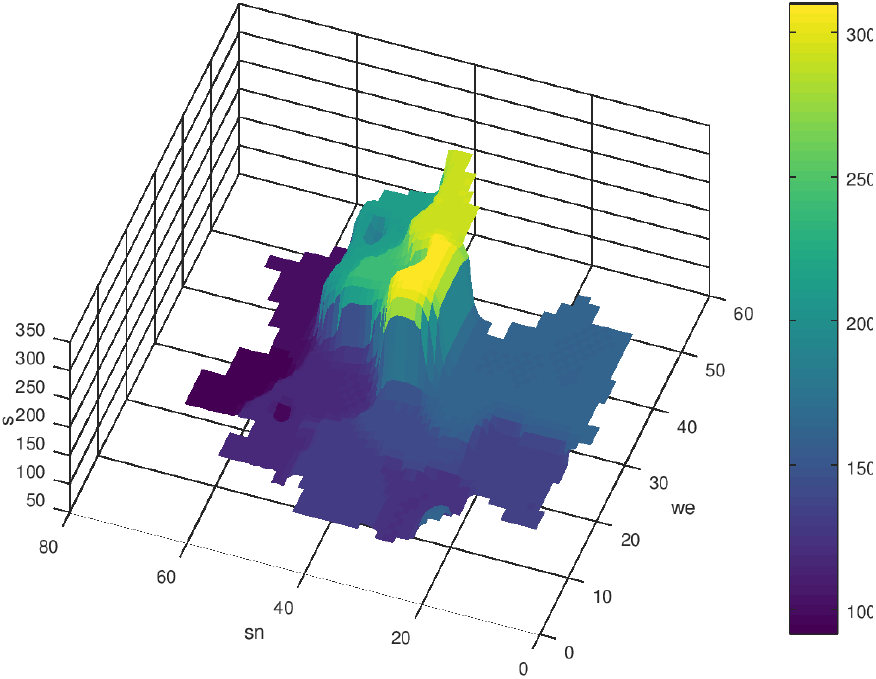
Initial distribution of *s, n* = 0, *q* = 0

**Figure 6:**
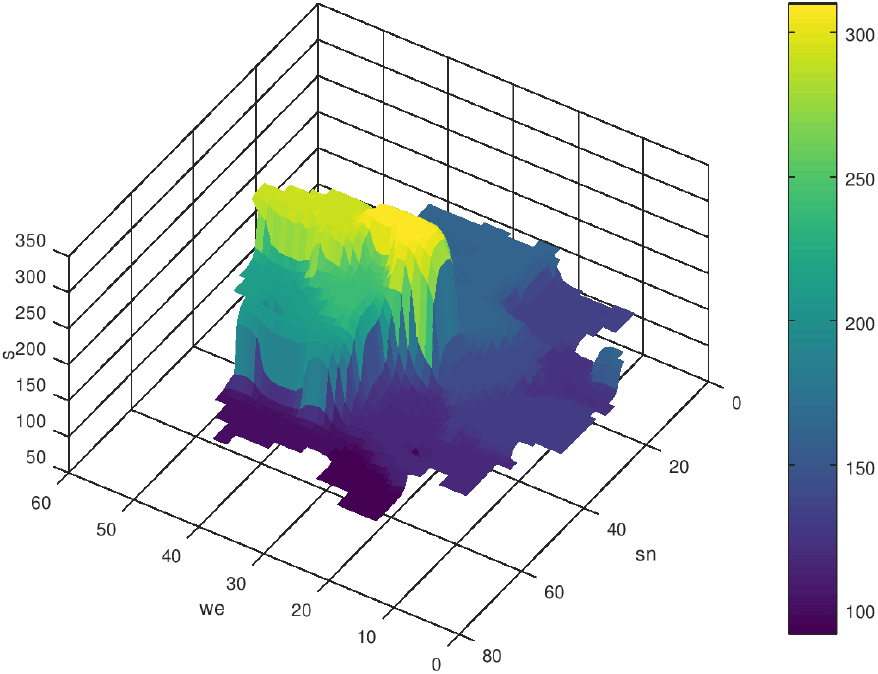
Initial distribution of *s, n* = 0, *q* = 0

**Figure 7:**
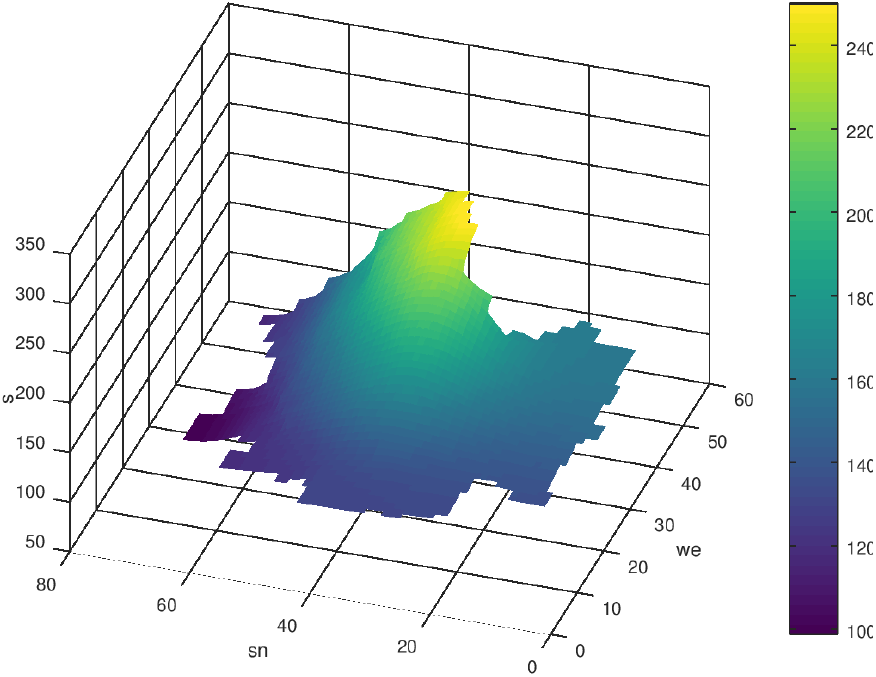
*s* after 100 time-steps, *q* = 0

**Figure 8:**
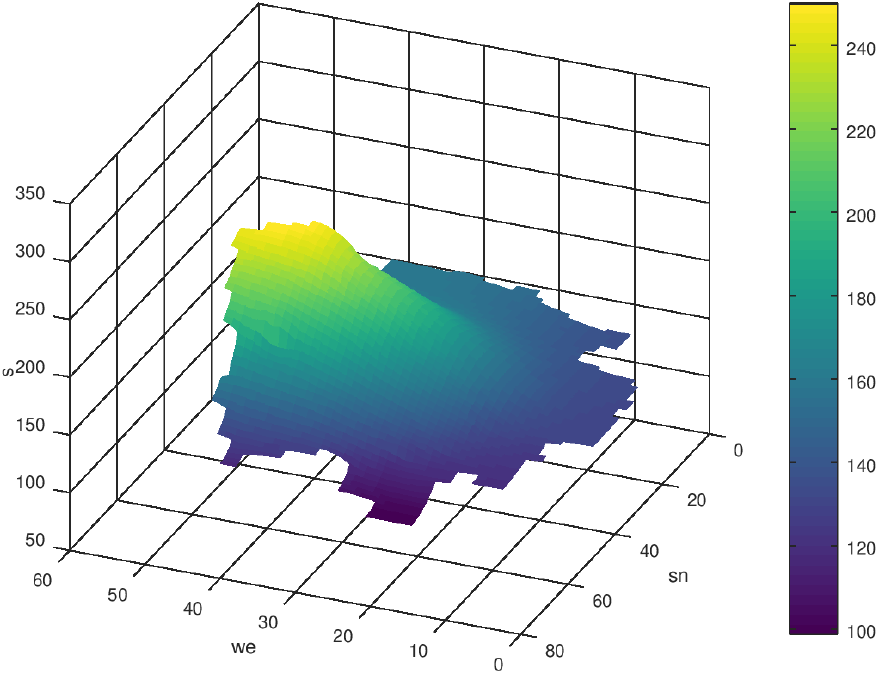
*s* after 100 time-steps, *q* = 0

**Figure 9:**
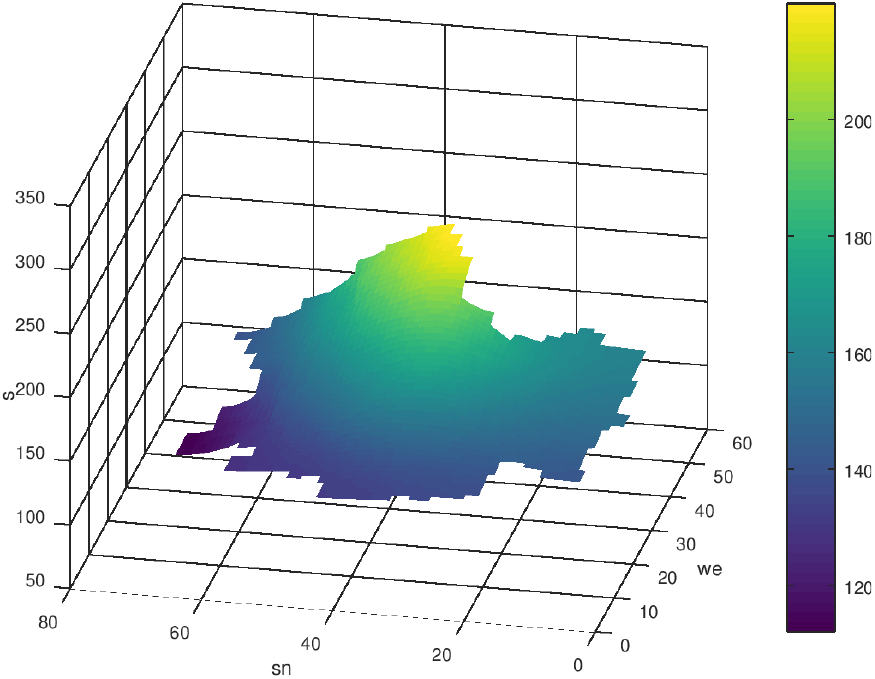
*s* after 200 time-steps, *q* = 0

**Figure 10:**
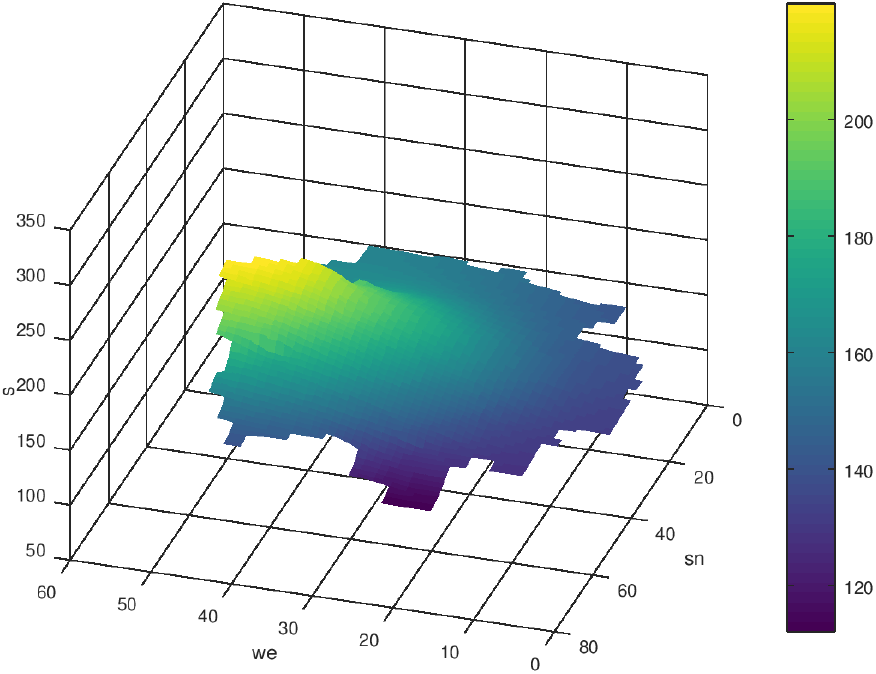
*s* after 200 time-steps, *q* = 0

With the parameters *α, β* and *γ* of the boundary condition (3) it’s possible to describe several situations at the borders of the boundary Γ of Ω. *α* = 0, *β* = − *D* and *γ* ≠ 0 describes a flux through the border. In the next example such a scenario will be used to describe the way home of infected people from Austria to Bavaria. The boundary condition at the border crossing reads as

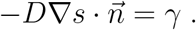

The initial state *s*_0_ was the same as in the example above. *γ >* 0 means an “inflow” of infected people, *γ <* 0 a loss of infected people (*γ* = 0 describes a closed border). In the following figures 11, 12 the case *γ* = 0 is compared to the case *γ* = 50 (*km/day*). At the south boundary of Bavaria one can observe the increase of *s* caused by the flux of *s* from Austria to Bavaria.

**Figure 11:**
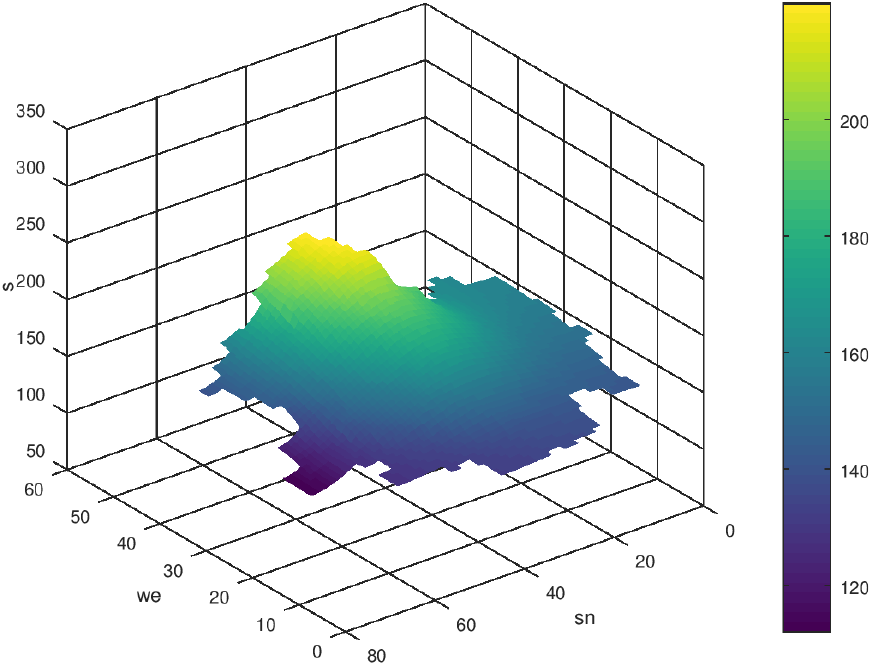
*s* after 200 time-steps, *γ* = 0*km/day, q* = 0

**Figure 12:**
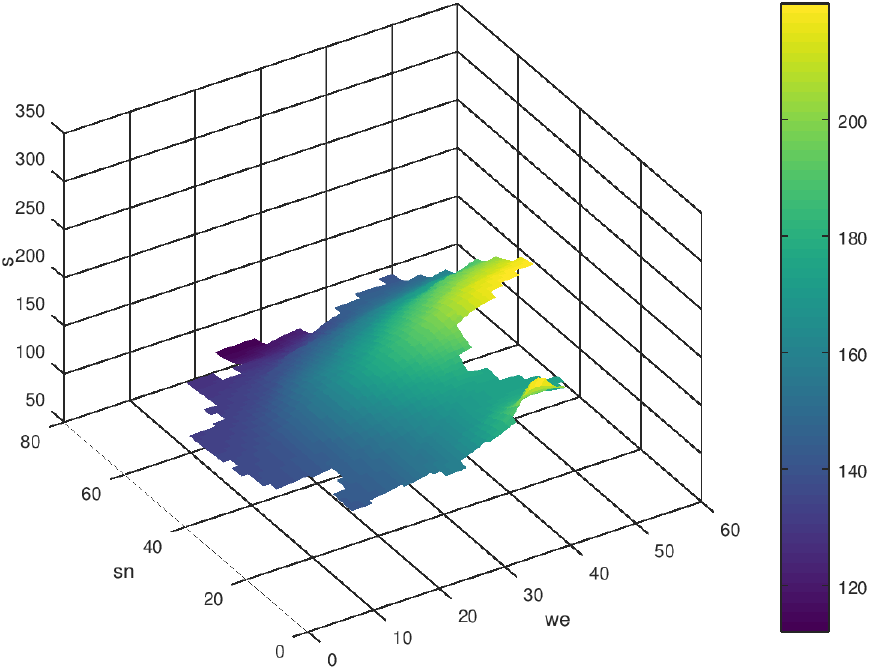
*s* after 200 time-steps, *γ* = 50*km/day, q* = 0

To get an idea of an appropriate approach of the source-sink term *q* let us consider the development of the 7-days incidence of three successive days in the federal states. The guessed values of the change of *s* per day are results of a linear regression. The change of *s* per day can be divede into a part coming from diffusion and another part coming from the local passing on the virus. We assume a 10% part coming from local transmission. The figures 13-16 show the simulation results with the assumptions for *q* after 10, 20, 30 and 50 time-steps (no border-crossing). The initial values are taken from table 1, i.e. the *s*-data of 12.01.2021.

**Figure 13:**
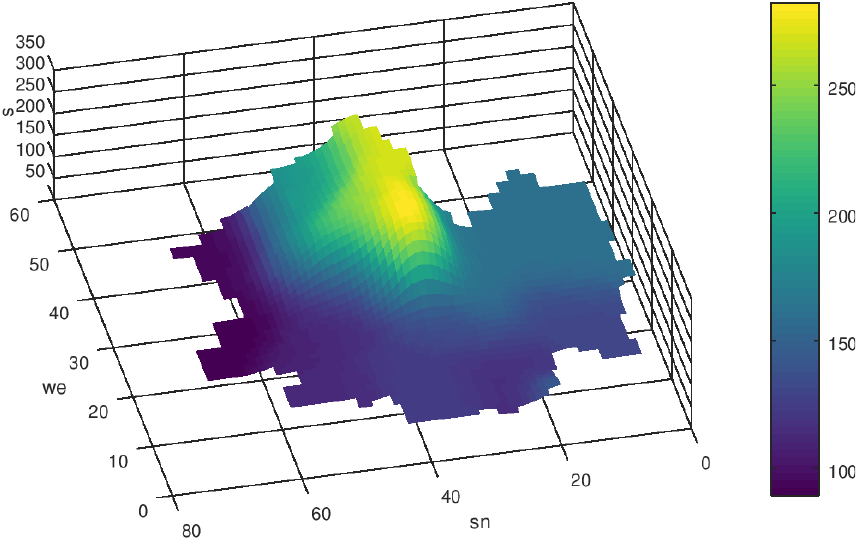
*s* after 10 time-steps, *q* ≠ 0, based on table 2

**Figure 14:**
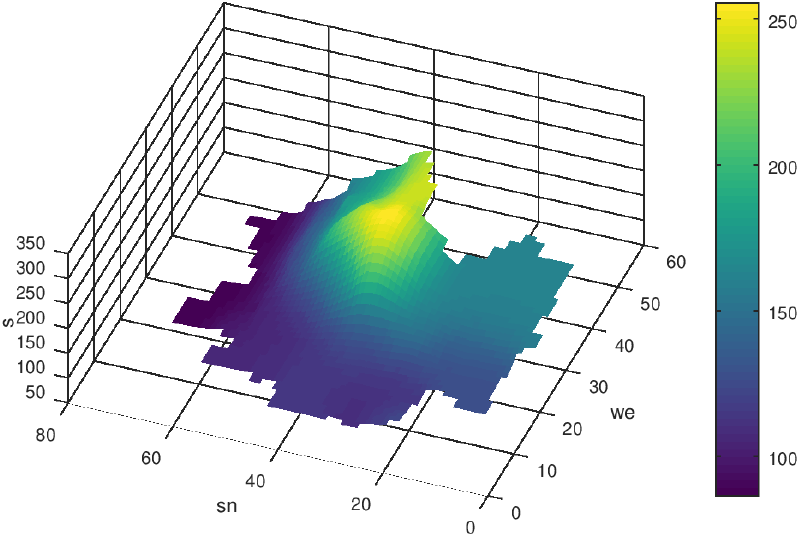
*s* after 20 time-steps, *q* ≠ 0, based on table 2

**Figure 15:**
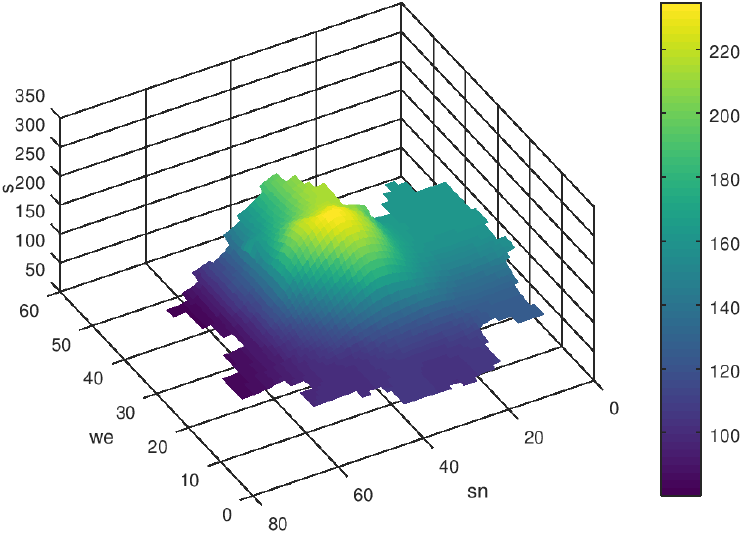
*s* after 30 time-steps, *q* ≠ 0, based on table 2

**Figure 16:**
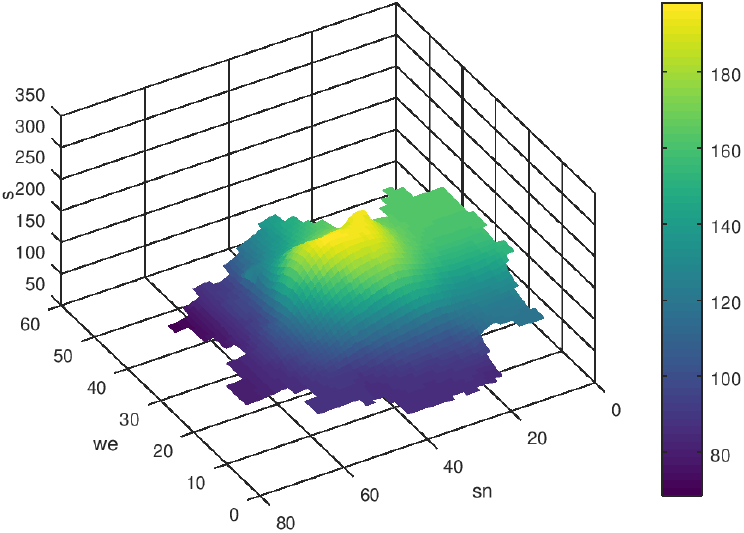
*s* after 50 time-steps, *q* ≠ 0, based on table 2

Because of the coarse approach of *q* it is advisable to update *q* based on the actual data, especially if the local pandemic progression is changing stiffly.

**Table 2:**
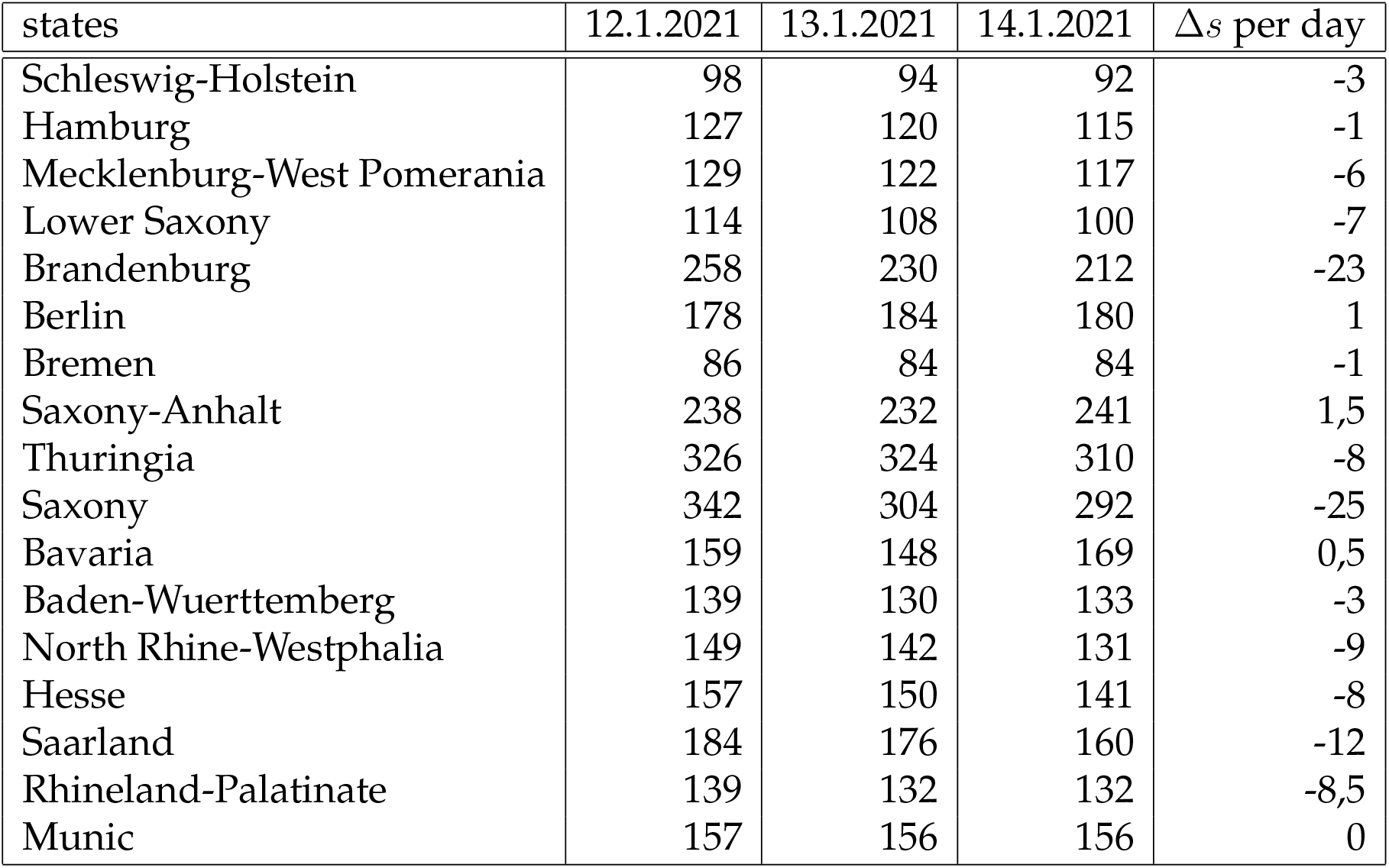
7-days incidence, guessed changed *s* per day [*/day*]

## 5 Discussion and Conclusion

The examples show the impact of diffusion effects on the propagation of the COVID-19 pandemic. It must be remarked that these processes are very slow compared to the virus transmission in a local hotspot cluster. But with the presented model is it possible to describe creeping processes which occur beside slack measures like holey lockdowns.

Especially the pandemic propagation in regions with high incidence gradients can be described with the discussed diffusion concept.

It would be interesting to continue the work with diffusion models especially the investigation of spreading events by a refined approach of *q* and the influence of border traffic. The presented model is qualified for such investigations. But it’s important to note that the diffusion modeling is only a small part of the understanding of the expansion of the SARS-CoV-2 virus.

At the end it should be noted that the distribution and propagation must be count back to the distribution and propagation of the infected people. Doing this, one can see, that the dense peaks of infected people are located in the metropolitan areas like Berlin, Munic or Hamburg. Figs. 17 and 18 show the distribution of the seven-days incidence and of the resulting infected people (by counting back using the people densities and the area of the federal states).

**Figure 17:**
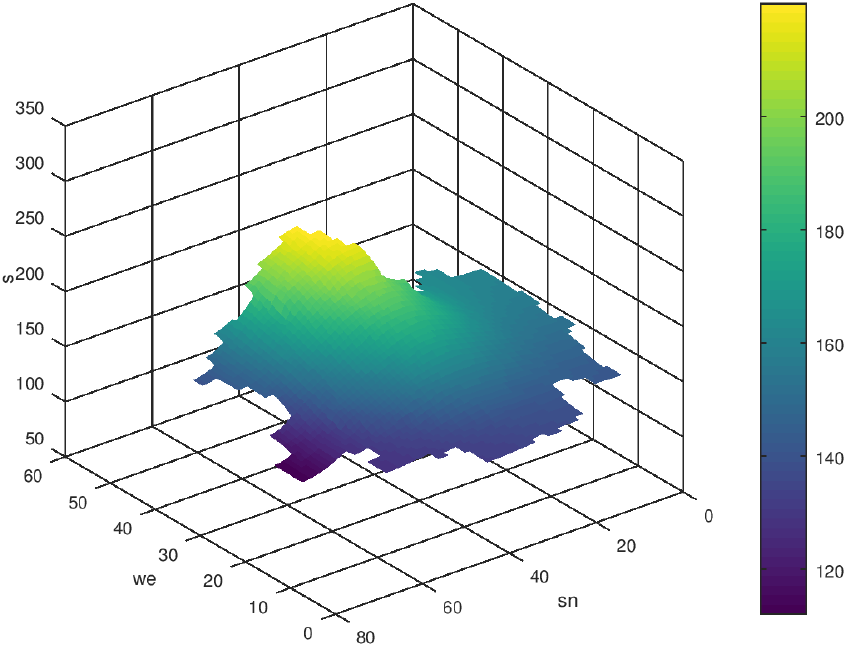
*s* after 200 time-steps, *q* ≠ 0

**Figure 18:**
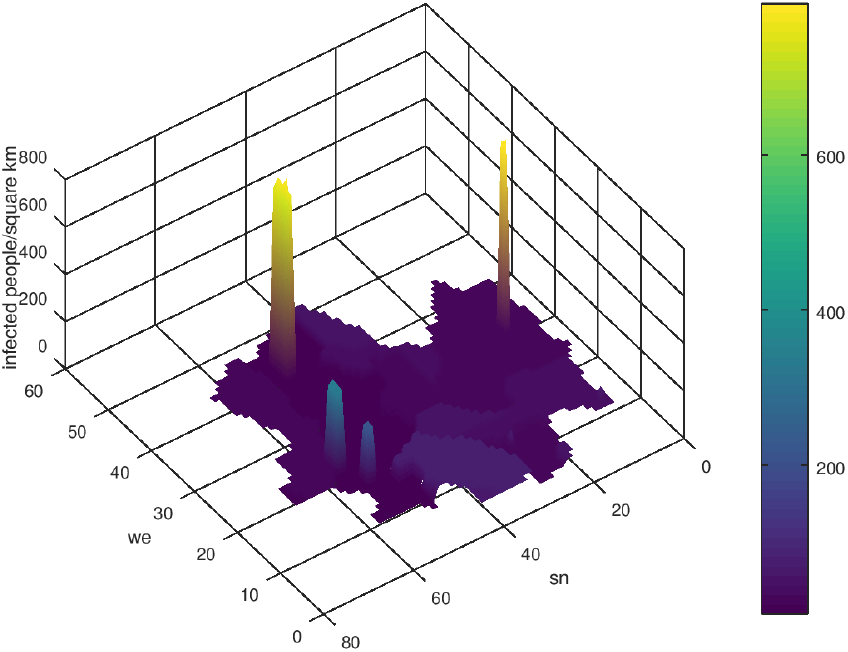
Infected people after 200 time-steps *q* ≠ 0

This research received no external funding.

## Data Availability

All used data are available on open websites.

https://www.rki.de/DE/Home/homepage_node.html

## Acknowledgements

Considering the topic of the pademic modeling I had some interesting exchange of ideas with my friends and colleagues F. Bechstedt, physicist of the Friedrich-Schiller University Jena, and Reinhold Schneider, mathematician of the Technical University Berlin. For this heartthanks.

## Footnotes

1 From gettyimages

